# Plant power? A systematic review of the effects of plant-based diets on people with mental illness

**DOI:** 10.1101/2020.09.28.20203026

**Authors:** Heather Catt, Jane Beenstock, Ummaz Nadeem, Adam Joiner

**Affiliations:** Department of Population Health, Health Services Research & Primary Care, University of Manchester, Manchester, UK, Lancashire and South Cumbria NHS Foundation Trust; Lancashire and South Cumbria NHS Foundation Trust, Lancaster University; Lancaster University; Lancashire and South Cumbria NHS Foundation Trust

**Author notes:** **Corresponding author** Heather Catt, Department of Population Health, Health Services Research & Primary Care University of Manchester, Stopford Building, Oxford Road, Manchester, M13 9PT.

**Keywords:** Mental health, plant-based diets, systematic review

## Abstract

**Objective:** There is increasing interest in plant-based diets in the general population and an increasing evidence base for the positive impact of plant-based diets on health outcomes for many chronic diseases. This systematic review aims to identify the effects of plant-based diets on people with mental health conditions.

**Methods:** A systematic review of intervention and observational studies. We conducted a systematic electronic search of MEDLINE (Ovid), EMBASE (Ovid), PsycINFO (ProQuest), British Nursing Index (ProQuest), CINAHL (EBSCO) and the Cochrane library to April 2019, with no date limits. We extracted data on outcomes and assessed the studies for bias using validated tools.

**Results:** We retrieved 588 studies. One study met the inclusion criteria with high risk of bias. The intervention was a plant-based diet for people with moderate to severe depression, without a control group. The study recruited 500 people, but recorded 66.8% attrition. Of the completers, 62% reported improvements in depressive symptoms, and 59% in anxiety symptoms. Completers lost 5.7lbs (2.6kg) during the trial and 15lb (6.8kg) at six month follow up.

**Conclusion:** There is not enough research to make conclusions about the effects of plant-based diets on people with mental health conditions. Given the evidence for positive effects of plant-based diets on physical health, further research is urgently required to understand the effects on people with mental health conditions. This will support the provision of advice and guidance for patients with mental illness who want to optimise their diet to improve their mental and physical health.

**PROSPERO registration:** CRD42019133440

## Introduction

Plant-based diets and vegetarian diets are terms used interchangeably for a group of diets which exclude or minimise the consumption of meat, including vegan diets which also exclude eggs and dairy. There is evidence that suggests that plant-based and vegetarian diets have beneficial effects on physical health, which include reduced risk of developing diabetes and cardiovascular disease, improved diabetic control and weight loss in patients with type 2 diabetes, and improved survival outcomes for people with colorectal cancer [1–7]. A recent systematic review found evidence for the positive impact of a plant-based diet on metabolic measures in health and disease [8]. This evidence has been translated into clinical and public health practice, with plant-based dietary patterns incorporated into guidelines for sustainable health and wellbeing [9–11], as well as Canadian and American guidelines for the management of type 2 diabetes [12,13].

The World Health Organisation predicted in 2001 that 450 million people were affected by a mental health condition globally [14]. A more recent review suggests that mental disorders and addictive disorders combined affect about one billion people globally [15]. Each year about 20% of the English population will experience a common mental disorder, yet perhaps only one in three will access help [16,17].

People with severe mental illness are more likely to have other chronic health conditions and die 15-20 years younger than the general population [18,19]. Those with mental health disorders experience higher rates of overweight and obesity than the general population due to a range of risk factors; medication, poor diet, high alcohol intake and inactive lifestyle [18]. Whilst not the only risk factor, medication can play a big part in weight gain: many medications used in psychiatry are associated with weight gain, particularly antipsychotics [20].

When people are started on antipsychotics, they tend to experience excessive and rapid weight gain [21–23]. The higher rates of abdominal obesity, metabolic syndrome, hypertriglyceridemia, diabetes and hypertension contribute to the life expectancy gap [24–27]. Whether or not plant-based diets can have positive health effects for people with severe and enduring mental illness is not known, but has been posited as worthy of exploration [28].

Whole (unprocessed) plant-based foods may be able to promote psychological wellbeing and reduce the risk of developing anxiety and depressive symptoms in the general population according to evidence from a range of study types [29–36]. However, a recently published systematic review highlights that the evidence for mental effects of a plant-based diet in the general population remains inconclusive [8]. A recent randomised controlled trial found that a healthful dietary pattern (plant foods with lean meats and low-fat dairy) could improve depressive symptoms for people with major depressive disorders, who were previously consuming an unhealthy diet [35]. This suggests that dietary interventions may support improved mental health outcomes for the population with a mental illness. Dietary interventions could be a low cost, public health intervention with potential benefits for mental health but also on physical health co-morbidities. As such, this is an important topic for consideration from both a clinical psychology and public health perspective.

Therefore, we conducted a systematic review of the literature to determine the benefits and risks of plant-based diets to people with mental illness and mental health conditions.

## Methods

### Systematic Search

We registered a review protocol with the International Prospective Register of Systematic Reviews (PROSPERO) database (CRD42016027656 http://www.crd.york.ac.uk/PROSPERO). We conducted a systematic electronic search of MEDLINE (Ovid), EMBASE (Ovid), PsycINFO (ProQuest), British Nursing Index (ProQuest), CINAHL (EBSCO) and the Cochrane library to April 2019, with no date limits. The search strategy included terms for plant-based diets and mental illness. Search words were plant-based diets or vegetarian or vegan and diet, and mental illness or mental health or mentally ill or mental problem or psychological disorder or psychiatric disorder or anxiety or depression or psychosis or schizophrenia or bipolar or personality disorder or mood or emotion. The Medline search strategy is shown in Table 1. This was amended for other databases and supplementary searches are shown in Supplementary Tables 1 to 5. Grey literature searches were conducted using keywords “vegetarian”, “vegan”, “plant-based diet” and “mental illness” and “mental health condition”.

**Table 1:**
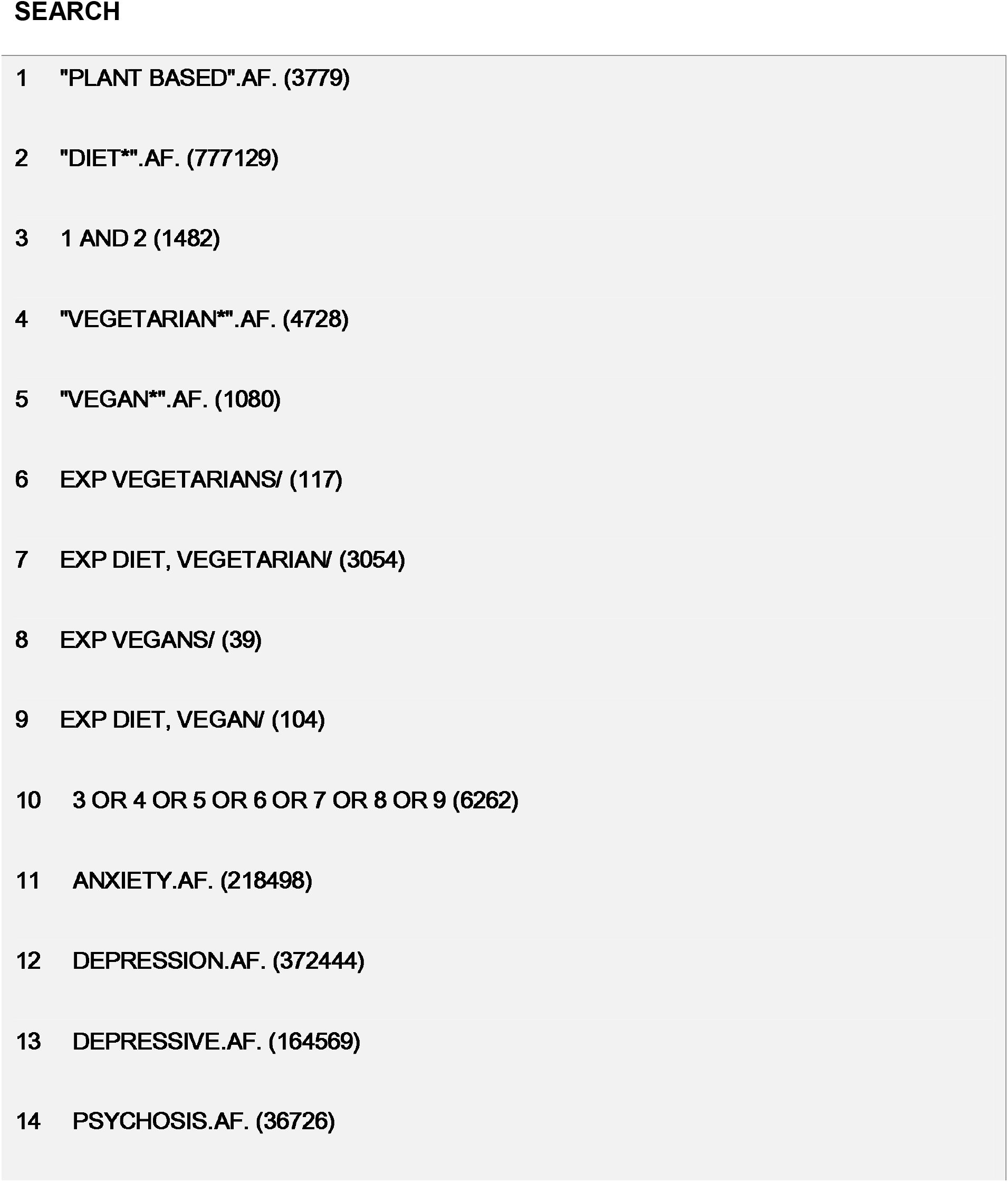
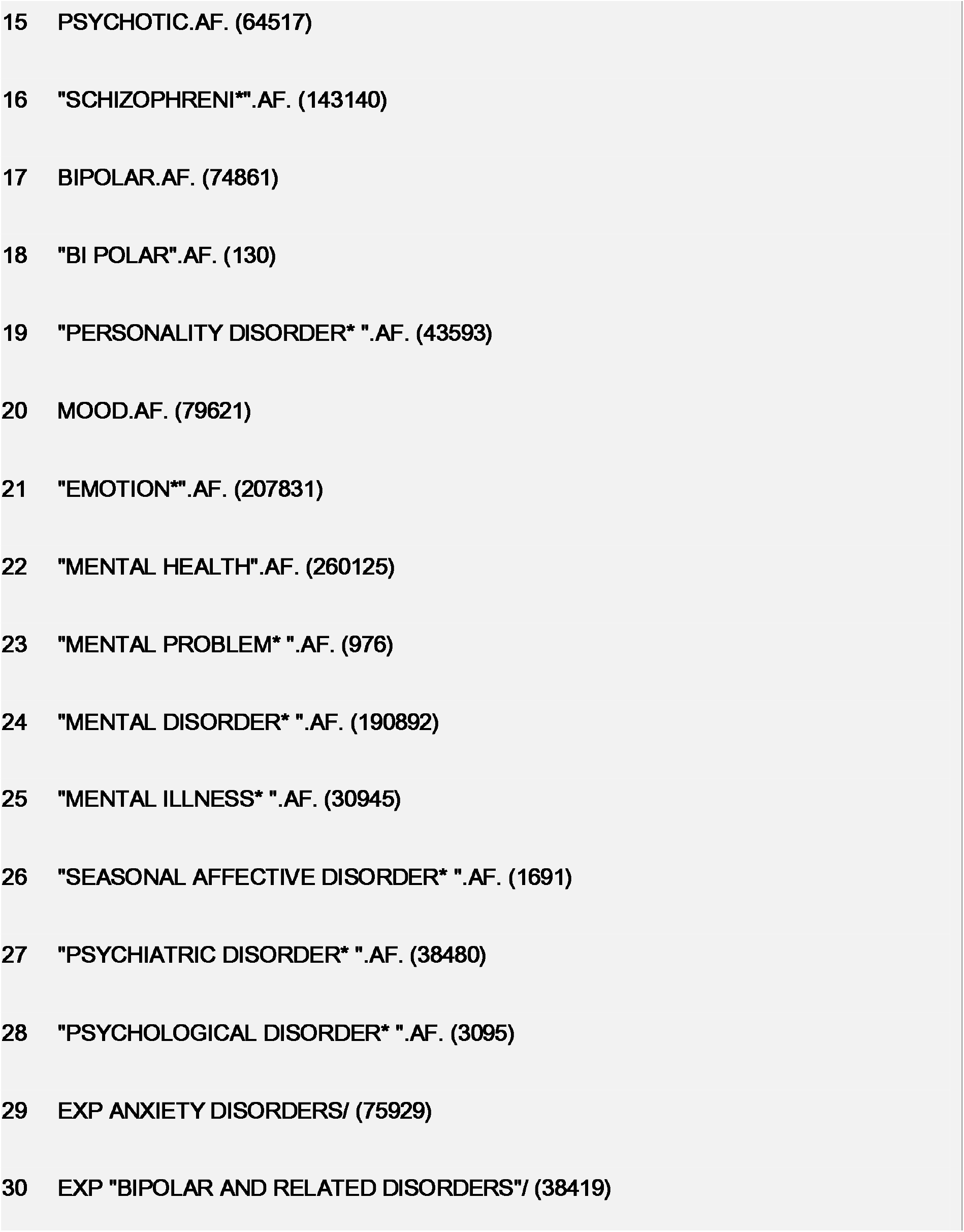
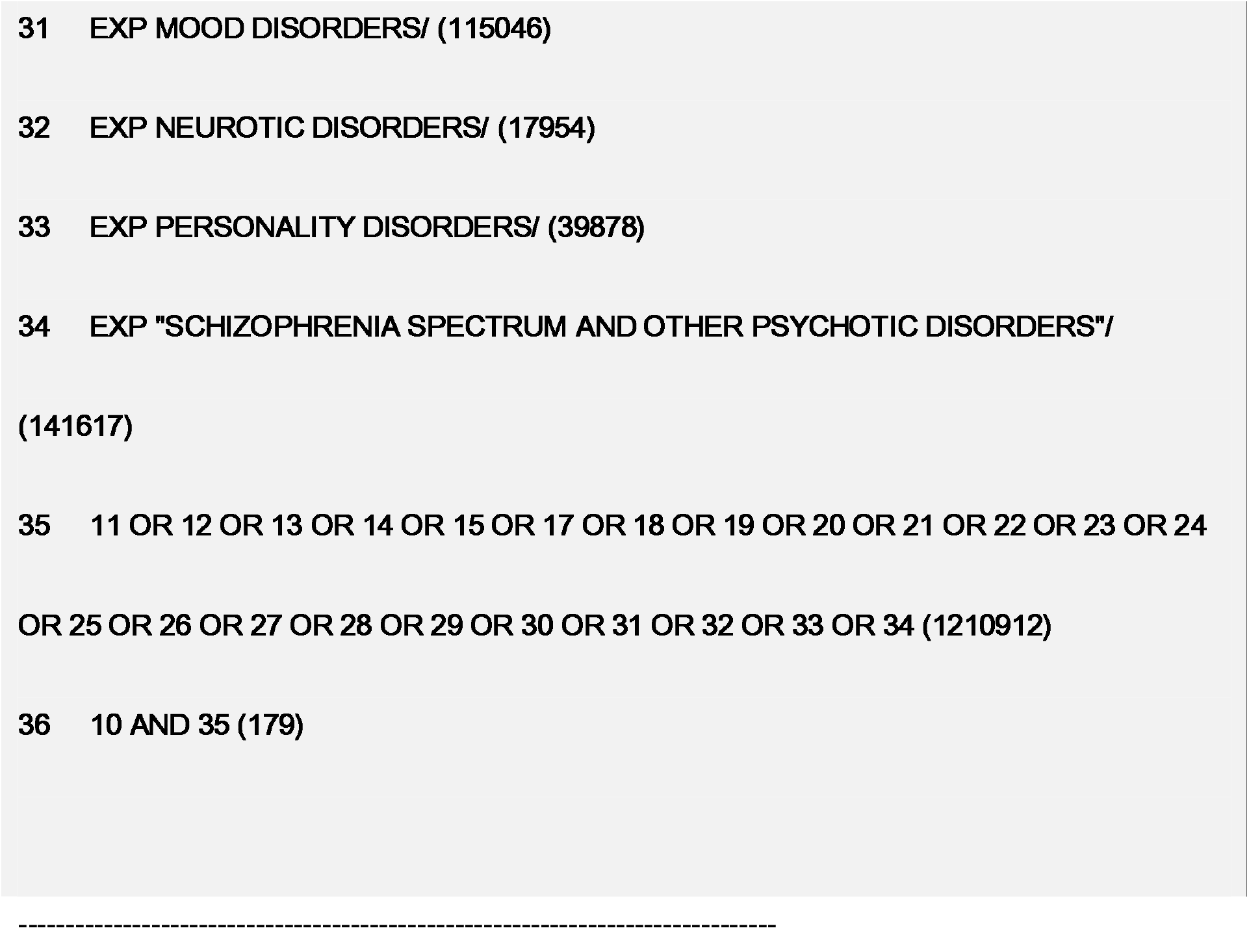
Ovid MEDLINE search strategy, 1946 to April 03, 2019>.

### Eligibility Criteria

RCTs, meta-analyses and systematic reviews to assess potential benefit of plant-based diets in people with mental illness were eligible. Observational studies were included to assess the benefits and harmful effects due to the likelihood of a limited number of RCTs. Studies would be included where they involved adults with neurotic and psychotic disorders (as diagnosed using recognised diagnostic criteria). Studies involving adolescents, and adults with learning disability, dementia, organic disorders and eating disorders were excluded. Studies had to be published as full text in English. Interventions of interest were plant-based diets, vegetarian diets and vegan diets. Comparator interventions of interest were usual diet, including omnivore diet, control diets and traditional calorie control diets. A broad range of outcomes were of interest including improvement in mental health symptoms, change in weight, metabolic status, disease markers, medicine doses and quality of life markers. Adherence outcomes were not included.

### Study Selection

Duplicates were removed after a complete list of studies was generated. An initial title screen was conducted by the librarian at Lancashire and South Cumbria NHS Foundation Trust. Two reviewers (JB and HC) independently assessed the remaining sample against eligibility criteria at the abstract screening stage and resolved discrepancies by discussion. Full copies were obtained of all potentially eligible studies and assessed against eligibility criteria by all three researchers (JB, HC and AJ). Discrepancies were resolved through discussion.

During the screening process, the decision was taken to widen the eligibility criteria to include interventions where plant-based diet was one component.

### Data Collection

A standardised form was developed to extract data from the included studies for assessment of study quality and evidence synthesis. Information was extracted on: study setting; study population, participant demographics and baseline characteristics; details of the intervention and control; study methodology; retention and completion rates; outcomes and times of measurement; suggested mechanisms of intervention action; information for the assessment of risk of bias. Two authors (JB and HC) extracted data independently and any discrepancies identified were resolved by discussion.

### Quality assessment

Risk of bias was to be assessed using a range of tools: the Cochrane risk of bias tool, the GRADE tool, the Newcastle-Ottawa scale and the ROBINS-I tool. Two reviewers (HC and AJ) independently assessed the risk of bias in included studies with disagreement resolved through discussion with a third reviewer (JB).

### Synthesis of Results and Analysis

Narrative synthesis of the results of the systematic review were planned due to the likely small number of diverse studies. The review was reported in line with the Preferred Reporting Items for Systematic Reviews and Meta-Analyses (PRISMA) statement (see Supplementary Table 6 for the PRISMA checklist) [37].

## Results

### Systematic search results

The database searches identified 850 records, 588 after duplicates were removed (Figure 1). No further results were identified from grey literature searches. Following title and abstract screen, 41 articles were included in the full review. All but one were ineligible, even with the widened inclusion criteria for the intervention.

**Figure 1:**
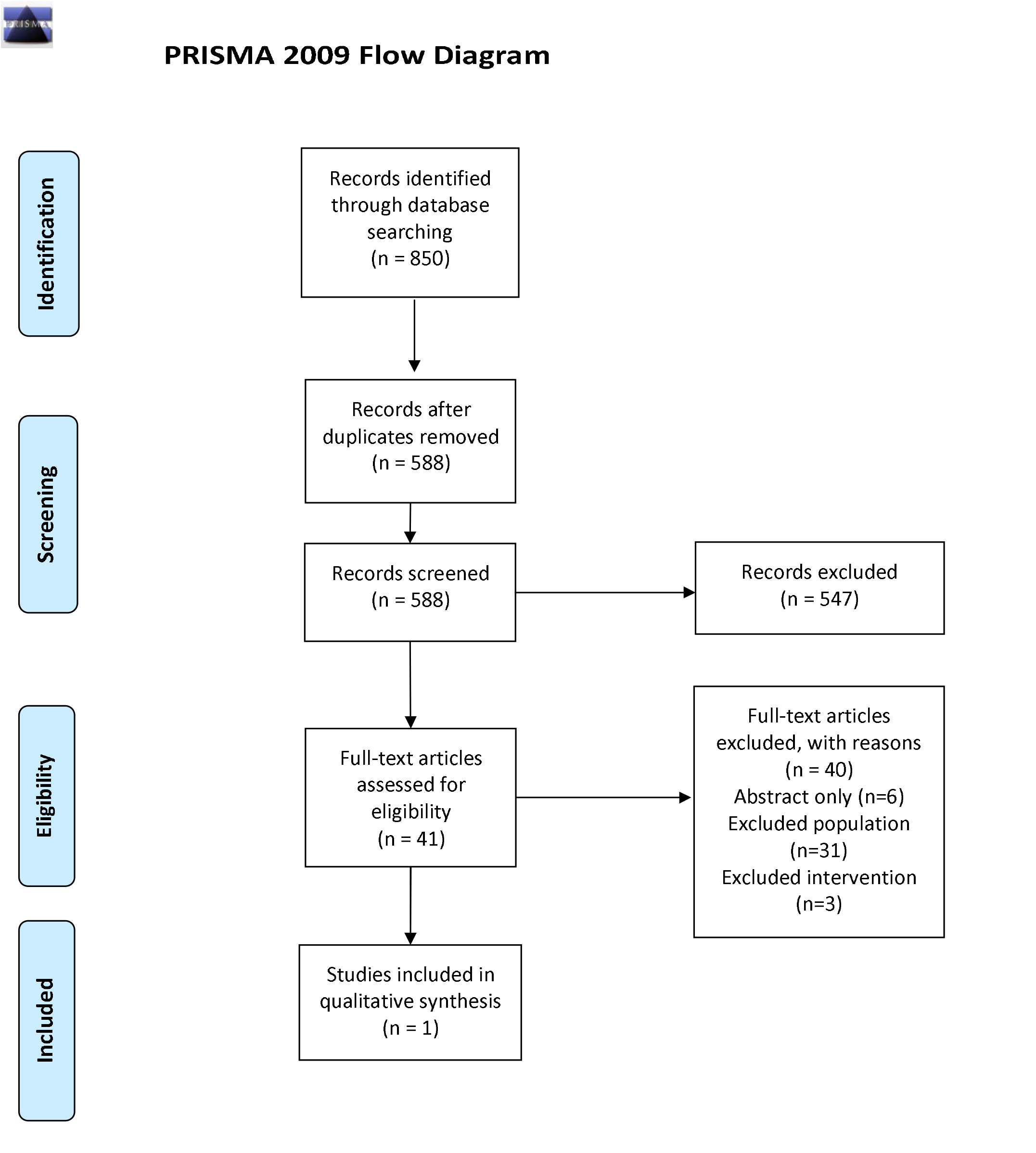
Preferred Reporting Items for Systematic Review and Meta-analyses (PRISMA) flow diagram. *From:* Moher D, Liberati A, Tetzlaff J, Altman DG, The PRISMA Group (2009). *P*referred *R*eporting *I*tems for *S*ystematic Reviews and *M*eta-*A*nalyses: The PRISMA Statement. PLoS Med 6(7): e1000097. doi:10.1371/journal.pmed1000097 For more information, visit www.prisma-statement.org.

The included study was of a 12 week, non-randomised, non-controlled diet and lifestyle intervention [38]. The extracted data is shown in Table 2. The study included 500 men and women with a diagnosis of chronic moderate to severe depression and anxiety. The diet component of the intervention was described as an anti-inflammatory plant-based diet with 70% raw foods, and no animal products. This was combined with a number of other components including juicing, exercise, mindfulness and environmental hygiene. Many outcomes were measured, but the key mental health outcomes were that 62% reported large improvement or complete remission of depression symptoms, 59% of anxiety symptoms. In terms of physical health, the key outcomes were average weight loss of 5.7lbs during the trial, increasing to 15.0lbs at six month follow up. However, 66.8% of the participations dropped out of the study, largely within the first two weeks and mostly because they found the diet and behaviour modification too rigorous. The results were reported as per protocol, rather than by intention to treat. The study was assessed to be of serious risk of bias.

**Table 2:** Characteristics of studies in the systematic review.

| Study | Study population | Patient demographics and baseline characteristics | Intervention | Control | Study methodology | Retention and completion rates | Outcomes and times of measurement | Results |
| --- | --- | --- | --- | --- | --- | --- | --- | --- |
| [38] | 500 men and women with diagnosis of chronic to severe depression and anxiety | Description of additional diagnoses for 166 who completed the project: 86 had moderate to major insomnia; 96 had moderate to major fatigue; 105 had moderate to major pain. | The study consisted of instruction on components of a healthy lifestyle: proper diet, juicing, detoxification, exercise, mindfulness and de-stressing techniques, environmental hygiene, and examination of beliefs and attitudes. Information on the prescribed intervention was given in weekly sessions of 2½ hours each, over a period of 12 weeks from the first meeting to the last.<br>Diet component was 70% raw food, whole-food plant based diet. Elimination of pro-inflammatory foods. Recipes provided and advice given to minimise costs of diet. | No control | Behaviour modification study to consider the impact of lifestyle and diet on individuals suffering from chronic moderate to severe depression and anxiety. Analysed on per protocol basis. | 334 (66.8%) dropped out. 90% in the first two weeks. 85% reported that the diet and behaviour modification was too rigorous. | Self-reported changes in physical, mental and emotional wellbeing (Likert scale)<br>Weight, blood pressure and body fat percentage were recorded at start and end of study<br>6 month survey conducted for feedback and other changes in health | 62% reported large improvement or complete remission of depression symptoms.<br>59% reported large improvement or complete remission of anxiety symptoms<br>64% of those affected by fatigue reported large improvement or complete remission<br>56% of those affected by major to moderate pain reported large improvement or complete remission<br>43% of those affected by insomnia reported large improvement or complete remission<br>The group's average systolic blood pressure fell 6.6 mmHg and the average diastolic blood pressure fell 3.5 mmHg.<br>The average weight loss among the group was 5.7 lbs and the average decrease in body fat percentage was 1.9%. At six month follow up, the average weight loss was 15.0lbs<br>At six month follow up, 78% reported that they were able to decrease or altogether eliminate antidepressant medications |

## Discussion

Given the increasing evidence base and guidance which advocates for plant-based diets to improve health outcomes in the general population [1–13], there is a concerning lack of evidence about the potential impact of plant-based diets on outcomes in people with mental health conditions. Our systematic review found just one study. While the results of this study found a plant-based diet (in a combined lifestyle intervention) had a positive impact on symptoms of people with major depressive disorder, the study was assessed as having serious risk of bias and the singular study means there is no data to synthesise.

There is some evidence of the impact of diet on the outcomes of people with mental health conditions. A randomised controlled trial found improvement in symptoms for people with major depressive disorder eating poor diets, who were subsequently helped to eat diets of whole, unprocessed, mostly plant-based foods, which included meat and dairy [39]. However, it is hard to know whether or not it was the whole plant foods which had the positive impact, or if including animal products had any bearing on this result. The diet recommended in this study aligns with the EAT-Lancet commission [11].

Although there is almost no research looking at the impact of exclusively plant-based diets on outcomes in people with mental ill health, there is some evidence for the impact of such diets on mental health and psychological wellbeing outcomes in the general population. Trials have suggested that different forms of plant-based diets (vegan and low fat, whole food diets) can lead to improvements in psychological wellbeing or positive impacts on mental health [31,36,40,41]. Further, the risk of developing depressive symptoms or major depressive episodes have been found to be inversely related to fruit and vegetable consumption and Mediterranean diets [29,30,42].

However, other evidence contradicts these findings and suggest that vegetarian diets may increase the risk of depressive symptoms demonstrating the lack of clarity even within the general population. Data from the Austrian health interview survey suggests that vegetarians are more likely to experience a mental health problem, than non-vegetarians [43]. Although questions have been raised about the potential for reverse causality or other confounding variables [44]. Some data suggests that vegetarian diets have no impact on mental health in Western populations [44], but can increase risk of anxiety or depressive symptoms in populations where vegetarianism is a consequence of economic restrictions rather than a positive health choice [45].

While it is generally accepted that diet does affect mental health [43], the current evidence is unable to provide clarity. Despite the lack of evidence, it remains plausible that diets of whole foods, which contain little or no processed foods, such as the Mediterranean diet, are likely to have a beneficial impact to reduce the risk of developing depressive symptoms, and contribute to recovering from depressive disorders. However, this needs further research, given the increasing evidence of the benefit of whole food plant-based diets on physical health [1–8], and the increase in people adopting vegan diets in developed countries. Britain’s vegan population has reportedly quadrupled between 2014 and 2019 to 600,000 [46]. Interest in vegan diets was recently demonstrated by analysis of google searches since 2004, which highlight a rapid growth in searches using the term “vegan” in Germany, the UK and the USA.[8]

What evidence that is available is not without its own limitations. Many studies which have looked at the effects of diet on mental health are relatively small, study a defined population (making generalisability difficult), and are open to many potential confounding factors, such a social norms, cultural influences, quality and type of foods consumed (both plant-based and animal-based).

There is significant variety in dietary patterns within groups such as ‘vegan’, ‘vegetarian’ and ‘omnivores’, making like-for-like comparisons difficult. It is possible to follow vegan and vegetarian diets which are composed of entirely processed foods with no fruit and vegetables, which would not be as healthy as a whole food plant based diet. Similarly, a diet which is composed of mostly whole plant-based foods with small amounts of lean meat is likely to be more beneficial for health than one without whole foods. These variations make comparisons of diets under broad headings difficult. There may be confounding effects from multiple other unknown sources also, such as pesticide use.

We must also be cautious using the current evidence base which focuses on depressive and anxiety symptoms of the general population, rather than those people suffering with mood disorders and anxiety disorders. There also seems to be a significant lack of research on the effects of dietary patterns on other mental disorders.

Given that there is increasing evidence for inflammatory processes in the aetiology of mental health conditions, [47,48] and that plant-based foods are considered as having a low dietary inflammatory index,[49,50] the interplay between plant-based diets, inflammation and mental illness requires further exploration. It is imperative to explore if intervening on a pro-inflammatory Western diet will have a positive impact on mental health.

## Conclusion

We have conducted a comprehensive systematic review to explore the impact of plant-based diets on mental illness, which is the first of its kind to our knowledge. A key strength of the study is the rigour of the methods. In addition, the results align with a recently published systematic review on the effects of plant-based diets on the body and the brain in the general population.[8] The authors of this review were able to demonstrate the impact of a plant based diet on metabolic measures in health and disease, but were unable to locate evidence for cognitive and mental effects. Our study differs as we were considering the research evidence for the population with mental illness, rather than the general population. Unsurprisingly, there is less evidence on the impact of a plant-based diet in this population and it is not possible to draw conclusions. Instead, this systematic review highlights this knowledge gap. It demonstrates the need for further research to explore this issue and elucidate the potentially important effects of diets on mental health. Clinically, the paucity of peer-reviewed publications on this topic indicates a concerning lack of evidence about how best to advice and guide patients with mental illness who want to optimise their diet to improve their mental and physical health. Observational research and randomised controlled trials are needed to explore the impact of plant-based diets (particularly wholefood plant-based diets) for people experiencing mental health conditions.

## Data Availability

No additional data is available.

## AUTHORSHIP

AJ, JB and UN developed the study concept. All authors contributed to the study design. Screening was performed by AJ, JB and HC. Data collection and synthesis were performed by HC. AJ, JB and HC drafted the paper. All authors approved the final version of the paper for submission.

### DATA ACCESS

All data is supplied within the document.

## ACKNOWLEDGEMENTS

Cath Harris, University of Central Lancashire for conducting the literature searches and the initial title screening.

---

**Supplementary table 1:**
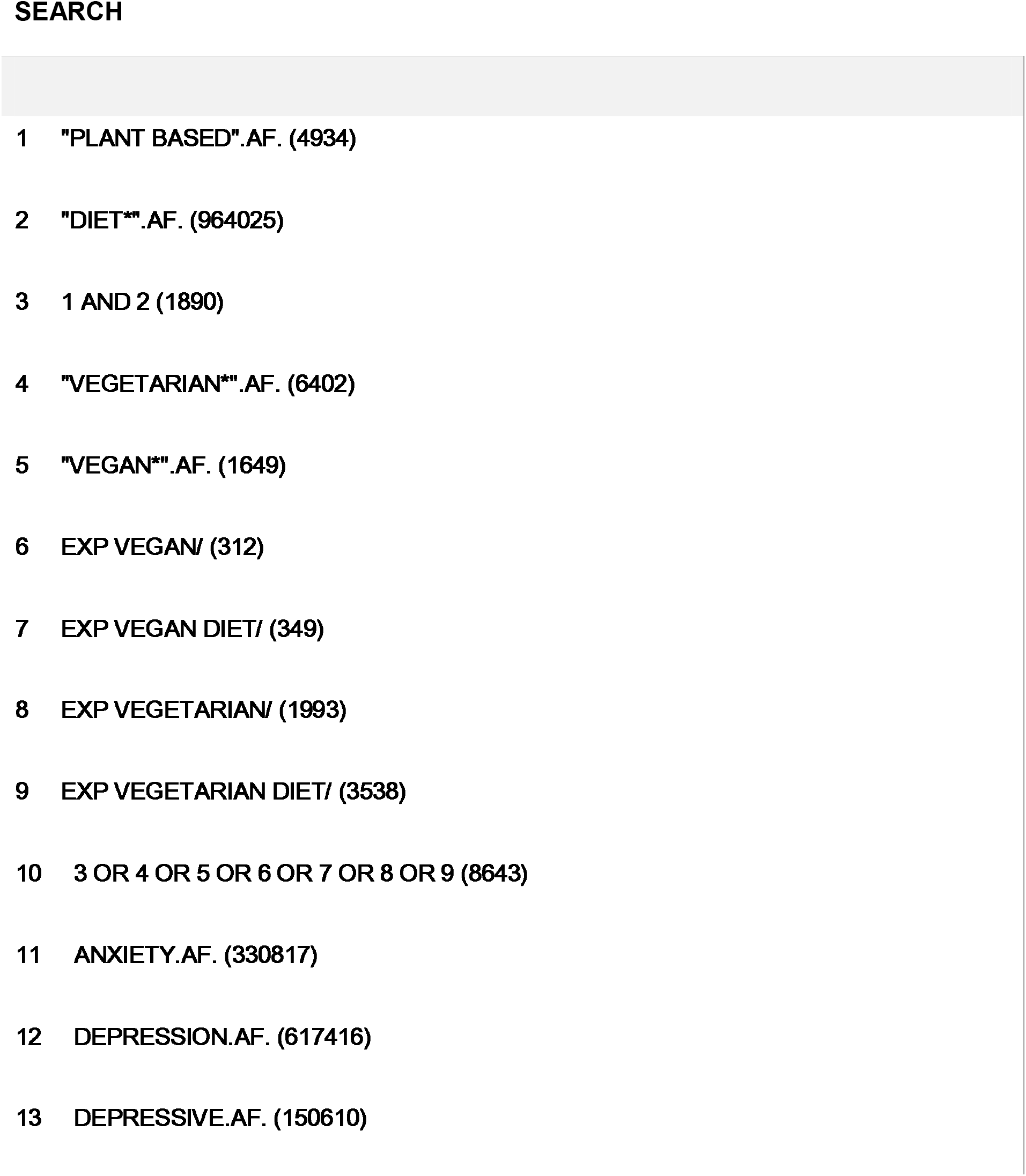
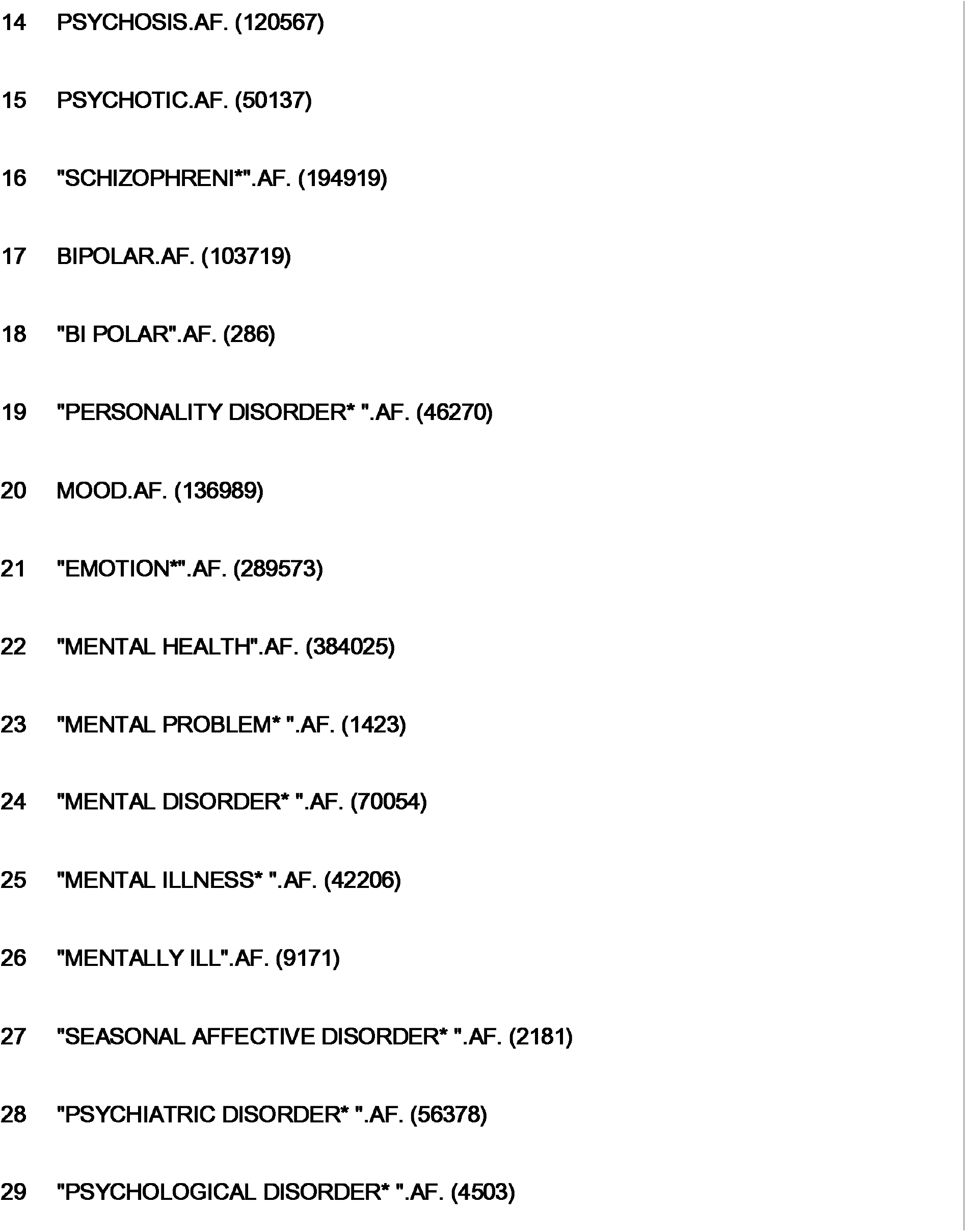
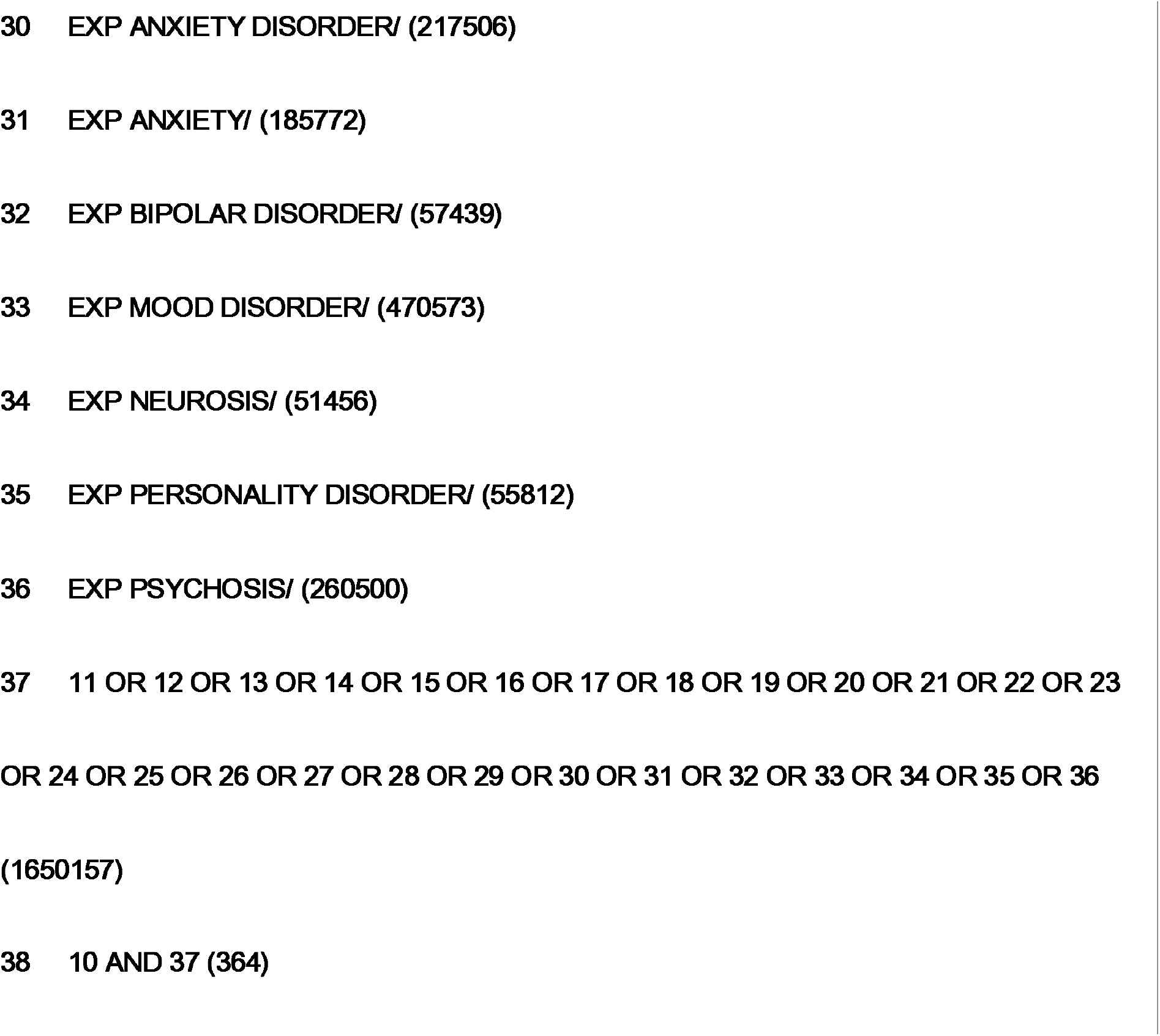
EMBASE search strategy, 1974 to 2019 April 03.

**Supplementary table 2:**
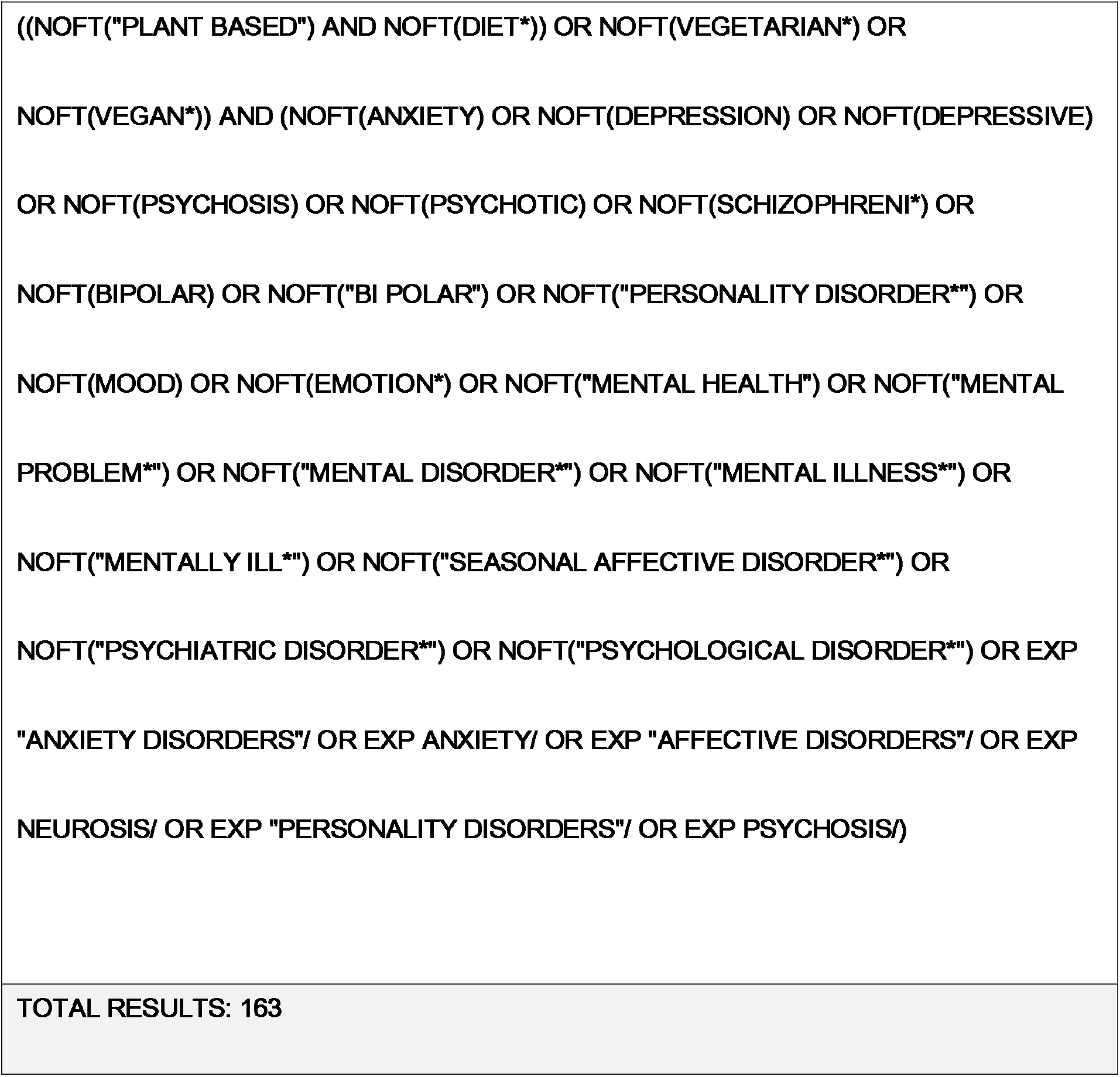
PsycINFO (ProQuest platform) search strategy.

**Supplementary table 3:**
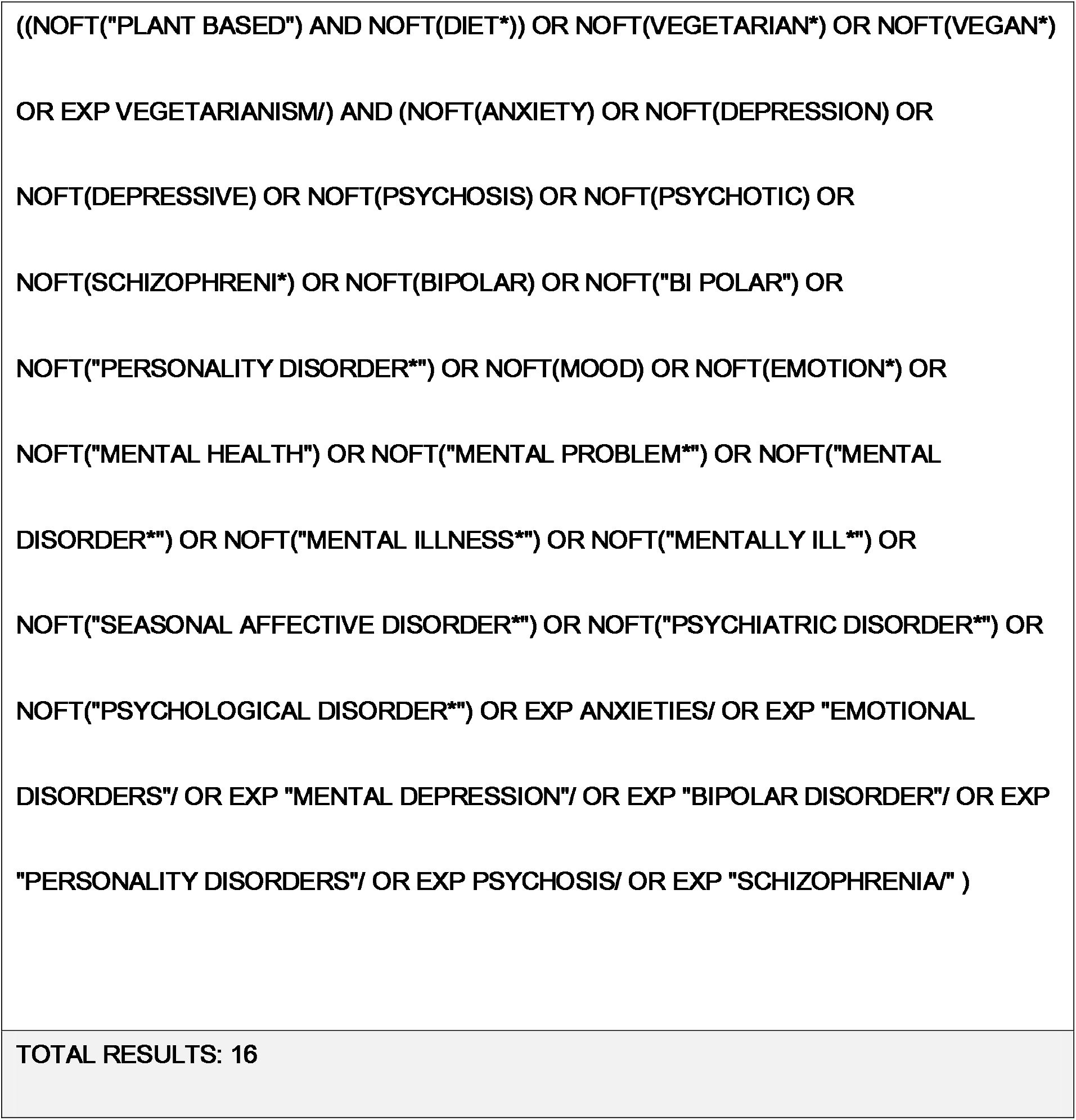
British Nursing Index (ProQuest platform) search strategy.

**Supplementary table 4:** CINAHL Plus with Full Test (EBSCO platform) search strategy.

|  |  |  |
| --- | --- | --- |
| S34 | S7 AND S33 | 128 |
| S33 | S8 OR S9 OR S10 OR S11 OR S12 OR S13 OR S14 OR S15 OR S16 OR S17<br>OR S18 OR S19 OR S20 OR S21 OR S22 OR S23 OR S24 OR S25 OR S26<br>OR S27 OR S28 OR S29 OR S30 OR S31 OR S32 | 505,107 |
| S32 | (MH "Psychotic Disorders+") | 104,741 |
| S31 | (MH "Personality Disorders+") | 12,205 |
| S30 | (MH "Neurotic Disorders+") | 134,582 |
| S29 | (MH "Affective Disorders+") | 101,352 |
| S28 | (MH "Bipolar Disorder+") | 10,256 |
| S27 | (MH "Anxiety Disorders+") | 38,689 |
| S26 | "psychological disorder*" | 951 |
| S25 | "psychiatric disorder*" | 8,832 |
| S24 | "seasonal affective disorder*" | 610 |
| S23 | "mentally ill*" | 4,778 |
| S22 | "mental illness*" | 19,231 |
| S21 | "mental disorder*" | 60,701 |
| S20 | "mental problem*" | 326 |
| S19 | "mental health" | 119,259 |
| S18 | emotion* | 87,788 |
| S17 | mood | 22,129 |
| S16 | "personality disorder**" | 10,464 |
| S15 | "bi polar" | 37 |
| S14 | bipolar | 15,840 |
| S13 | schizophreni* | 27,544 |
| S12 | psychotic | 14,659 |
| S11 | psychosis | 10,695 |
| S10 | depressive | 34,692 |
| S9 | depression | 141,247 |
| S8 | anxiety | 80,569 |
| S7 | S3 OR S4 OR S5 OR S6 | 6,247 |
| S6 | (MH "Vegetarianism") | 5,064 |
| S5 | vegan* | 1,145 |
| S4 | vegetarian* | 5,633 |
| S3 | S1 AND S2 | 700 |
| S2 | diet* | 185,267 |
| S1 | "plant based" | 1,083 |

**Supplementary table 5:**
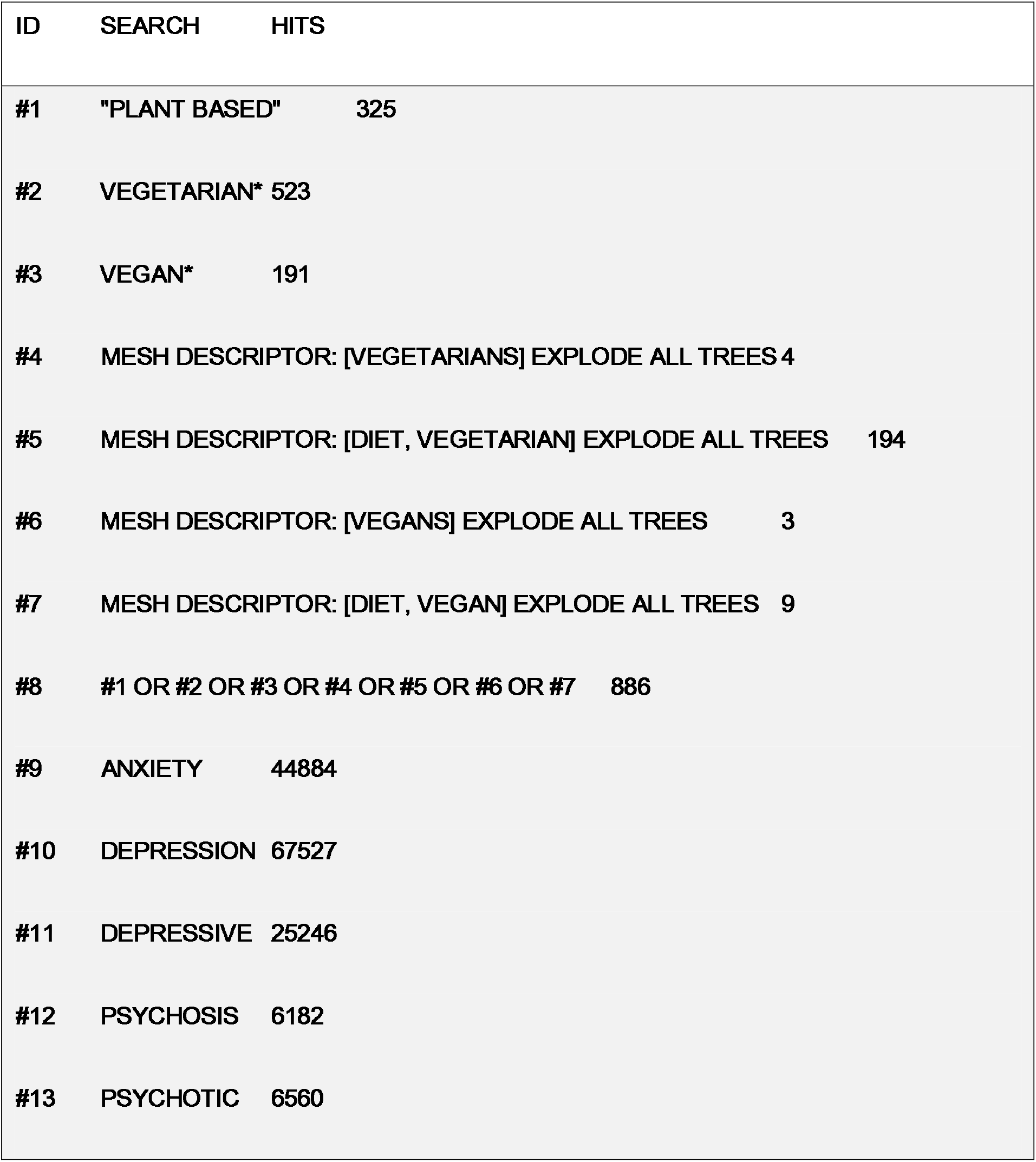
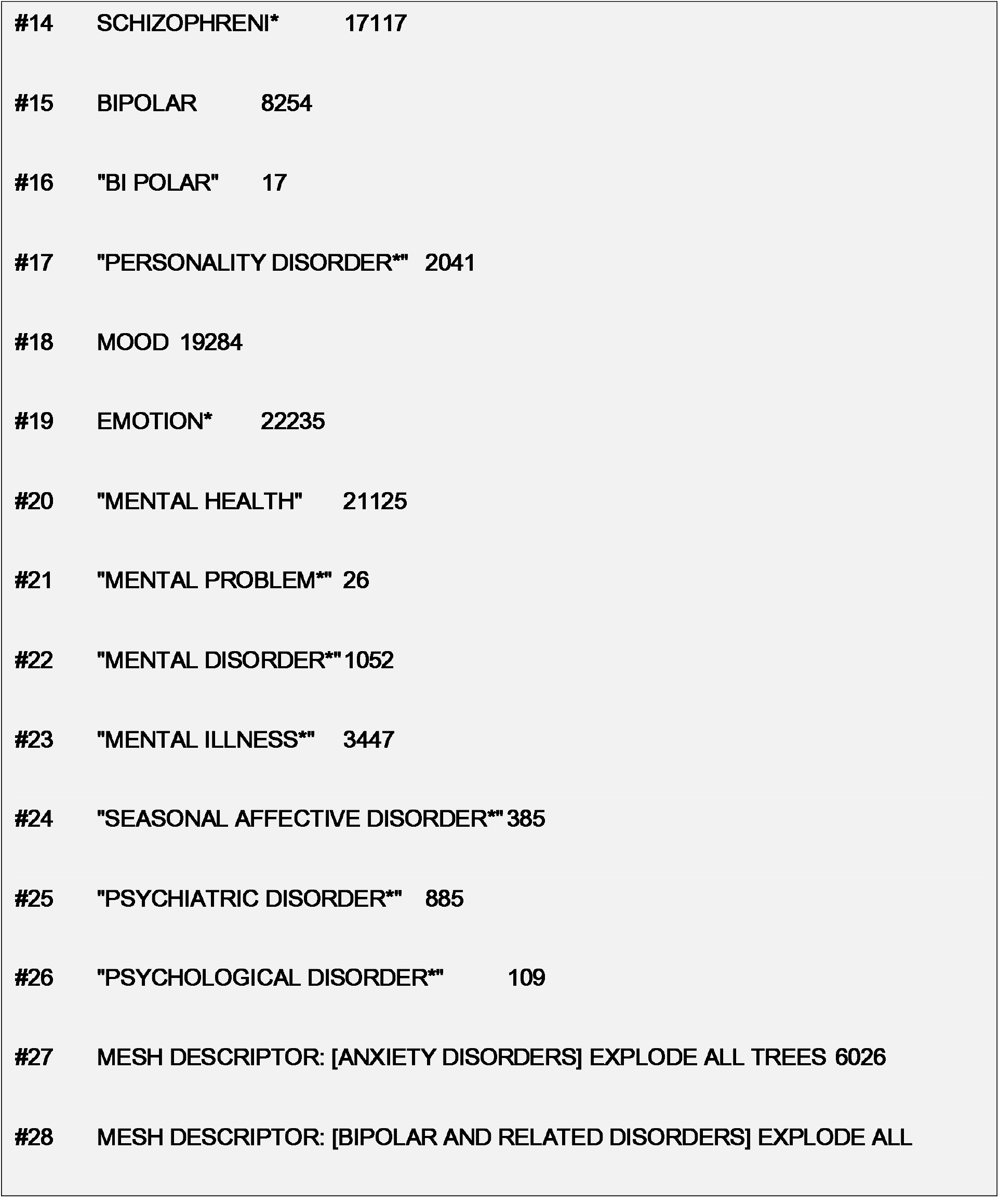
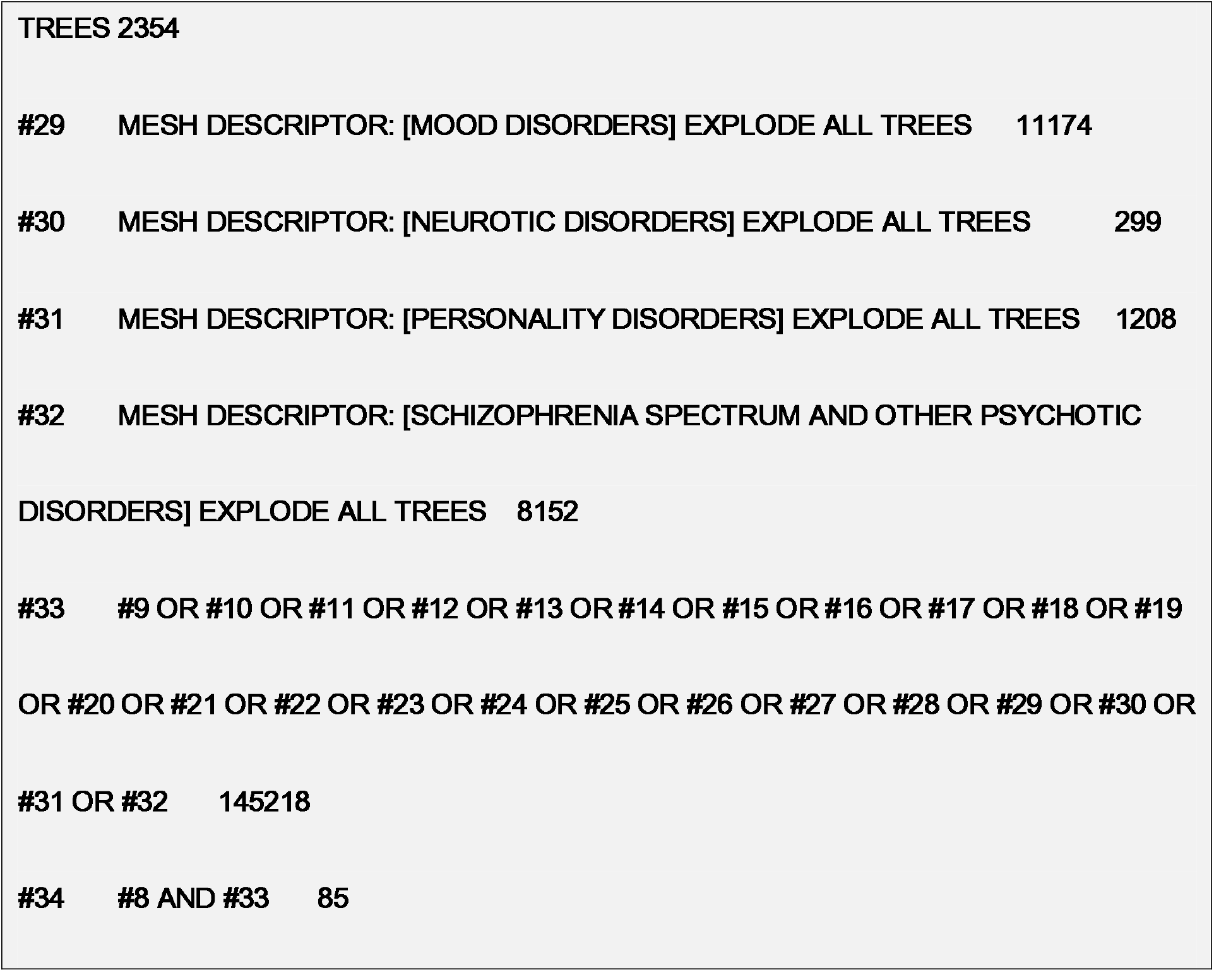
Cochrane library search strategy.

**Supplementary table 6:** Preferred Reporting Items for Systematic Review and Meta-analyses (PRISMA) checklist.

| Section/topic | # | Checklist item | Reported on page # |
| --- | --- | --- | --- |
| **TITLE** |  |  |  |
| Title | 1 | Identify the report as a systematic review, meta-analysis, or both. | 1 |
| **ABSTRACT** |  |  |  |
| Structured summary | 2 | Provide a structured summary including, as applicable: background; objectives; data sources; study eligibility criteria, participants, and interventions; study appraisal and synthesis methods; results; limitations; conclusions and implications of key findings; systematic review registration number. | 2 |
| **INTRODUCTION** |  |  |  |
| Rationale | 3 | Describe the rationale for the review in the context of what is already known. | 4-6 |
| Objectives | 4 | Provide an explicit statement of questions being addressed with reference to participants, interventions, comparisons, outcomes, and study design (PICOS). | 6 |
| **METHODS** |  |  |  |
| Protocol and registration | 5 | Indicate if a review protocol exists, if and where it can be accessed (e.g., Web address), and, if available, provide registration information including registration number. | 6 |
| Eligibility criteria | 6 | Specify study characteristics (e.g., PICOS, length of follow-up) and report characteristics (e.g., years considered, language, publication status) used as criteria for eligibility, giving rationale. | 7 |
| Information sources | 7 | Describe all information sources (e.g., databases with dates of coverage, contact with study authors to identify additional studies) in the search and date last searched. | 6-7 |
| Search | 8 | Present full electronic search strategy for at least one database, including any limits used, such that it could be repeated. | 30 |
| Study selection | 9 | State the process for selecting studies (i.e., screening, eligibility, included in systematic review, and, if applicable, included in the meta-analysis). | 8 |
| Data collection process | 10 | Describe method of data extraction from reports (e.g., piloted forms, independently, in duplicate) and any processes for obtaining and confirming data from investigators. | 8 |
| Data items | 11 | List and define all variables for which data were sought (e.g., PICOS, funding sources) and any assumptions and simplifications made. | 8 |
| Risk of bias in individual studies | 12 | Describe methods used for assessing risk of bias of individual studies (including specification of whether this was done at the study or outcome level), and how this information is to be used in any data synthesis. | 9 |
| Summary measures | 13 | State the principal summary measures (e.g., risk ratio, difference in means). | 9 |
| Synthesis of results | 14 | Describe the methods of handling data and combining results of studies, if done, including measures of consistency (e.g., $I^2$) for each meta-analysis. | 9 |

